# Development of an Australian Clinical Practice Guideline on methylenedioxymethamphetamine (MDMA)-assisted Psychotherapy for Post-traumatic Stress Disorder

**DOI:** 10.1101/2025.02.18.25322184

**Authors:** Alene Sze Jing Yong, Sue E Brennan, Suzie Bratuskins, Aimée Freeburn, Gillinder Bedi, Rimona Burke, Terry Haines, Mary Hollick, Kimberley Ann Jones, Andrew J Lawrence, Yong Yi Lee, Alexander C McFarlane, Stephen Parker, Nicholas Procter, Liam Spicer, Andrew A. Somogyi, Simon Stafrace, Stacey Watts, Clare Walton, Kay Wilson, J Simon Bell

## Abstract

**Introduction:** Despite recent clinical and research interest, medical use of psychedelics has not been legalised in most jurisdictions. The Australian Therapeutic Goods Administration rescheduled methylenedioxymethamphetamine (MDMA) in July 2023 to permit authorised prescribing of MDMA for Post-traumatic Stress Disorder (PTSD) outside of the clinical trial setting.

**Objective:** This manuscript describes the development of the Australian Clinical Practice Guideline on MDMA-assisted psychotherapy (MDMA-AP) for PTSD.

**Methods:** The Guideline will be developed using the Grading of Recommendations, Assessment, Development, and Evaluations (GRADE) process. The Guideline will consider the benefits and harms of MDMA-AP compared to other treatments in people with PTSD. High quality systematic reviews identified via an overview of systematic reviews will be used as index and supplementary reviews. Using the GRADE Evidence-to-Decision framework, the multidisciplinary Guideline Development Group (GDG) will consider treatment benefits and harms, certainty of evidence, patient preferences and values, resources, equity, acceptability and feasibility. The GDG will be supported by a Stakeholder Group, Expert Group, Conflict of Interest Oversight Committee, and Evidence Review Team. The Guideline will be developed using an integrated knowledge translation approach, emphasising the co-production of knowledge through active participation and shared decision-making with end-users.

**Conclusion:** The Guideline will be published on the digital platform MAGICapp and disseminated in peer-reviewed publications, professional conferences and via specific stakeholder groups. A Companion Guide will be developed for people living with PTSD and their carers, family members, and supports.

## 1. Introduction

Medical use of psychedelics has not been legalised in most countries. Canada, Israel and Switzerland restrict access to supervised use of psychedelics for compassionate reasons (Mocanu et al., 2022; Oehen & Gasser, 2022). In the US, the therapeutic use of psychedelics is legalised in some states, such as Oregon and Colorado (Psychedelic Alpha; Siegel et al., 2023). However, there is no approved psychedelic-containing product because the new drug application for methylenedioxymethamphetamine (MDMA)-assisted psychotherapy (MDMA-AP) was rejected by the US Food and Drug Administration (FDA) in August 2024, with the FDA citing the need for more research prior to approval (Lykos Therapeutics, 2024b).

In July 2023, the Australian Therapeutic Goods Administration rescheduled MDMA and psilocybin from being prohibited drugs (Schedule 9) to being controlled drugs (Schedule 8) (Therapeutic Goods Administration, 2023). This rescheduling permitted authorised psychiatrists to prescribe MDMA and psilocybin for post-traumatic stress disorder (PTSD) and treatment-resistant depression respectively, outside of the clinical trial setting. To be an authorised prescriber, a psychiatrist is required to follow a clinical treatment protocol and obtain approval from a human research ethics committee (Therapeutic Goods Administration, 2024). Following the rescheduling, the Royal Australian and New Zealand College of Psychiatrists (RANZCP) published a clinical memorandum for psychiatrists about the therapeutic use of MDMA and psilocybin (Royal Australian and New Zealand College of Psychiatrists, 2023). The memorandum recommended that MDMA and psilocybin in conjunction with psychotherapy should be reserved for those who have tried all other established evidence-based treatments without lasting success.

Both international and local stakeholders reported mixed perspectives toward psychedelic use for mental health conditions (Barnett et al., 2022; Barnett et al., 2018; Kucsera et al., 2023; Kunstler et al., 2023; Page et al., 2021). In the US, healthcare professionals, including psychiatrists, psychologists, therapists, and social workers, reported being unprepared to discuss with patients the medical use of psychedelics due to concerns about the lack of trained providers, logistical challenges, safety issues, potential psychiatric risks, and managing patients with contraindications (Barnett et al., 2022; Kucsera et al., 2023).

Australian community surveys have shown indecision or indifference towards the use and approval of psychedelics for treating mental health conditions (Kunstler et al., 2022; Nadeem et al., 2024). However, those people with mental illness may have more positive attitudes (Nadeem et al., 2024). Half of the Australian psychiatrists surveyed did not consider themselves knowledgeable about psychedelic therapies and many were concerned about safety, access, and regulation (Berger & Fitzgerald, 2023). Similarly, a more recent survey of general practitioners, psychiatrists, and psychologists indicated positive attitudes towards psychedelic-assisted psychotherapy’s potential to improve outcomes, but highlighted ongoing concerns about safety, efficacy, and the scientific rigor of current research (Nadeem et al., nd).

In a context where patients have access to psychedelic-assisted treatment but no approved products available, development of guidelines can support clinicians to deliver appropriate care and patients to make informed decisions. This manuscript describes the development of an Australian clinical practice guideline for the use of MDMA-AP in PTSD (herein referred to as the Guideline). This protocol details in advance the methods that will be used in the development of the guideline, with the aim of ensuring a transparent process and minimising the risk of bias (Ford et al., 2022). To our knowledge, this will the first clinical practice guideline related to the use of MDMA-AP for PTSD.

## 2. Methods

### 2.1 Governance Structure

The Guideline will be developed using the Grading of Recommendations, Assessment, Development, and Evaluations (GRADE) process (The GRADE Working Group, 2024) in accordance with National Health and Medical Research Council (NHMRC) Guidelines for Guidelines and 2016 Standards for Guidelines (National Health and Medical Research Council, 2016). The intent to develop the Guideline was registered by NHMRC on 7 February 2024 and NHMRC appointed a Guideline liaison person. The Guideline will be developed by the Guideline Development Group (GDG) supported by a Stakeholder Group, Expert Group, Conflict of Interest (COI) Oversight Committee, and Evidence Review Team (ERT) (Figure 1).

**Figure 1.**
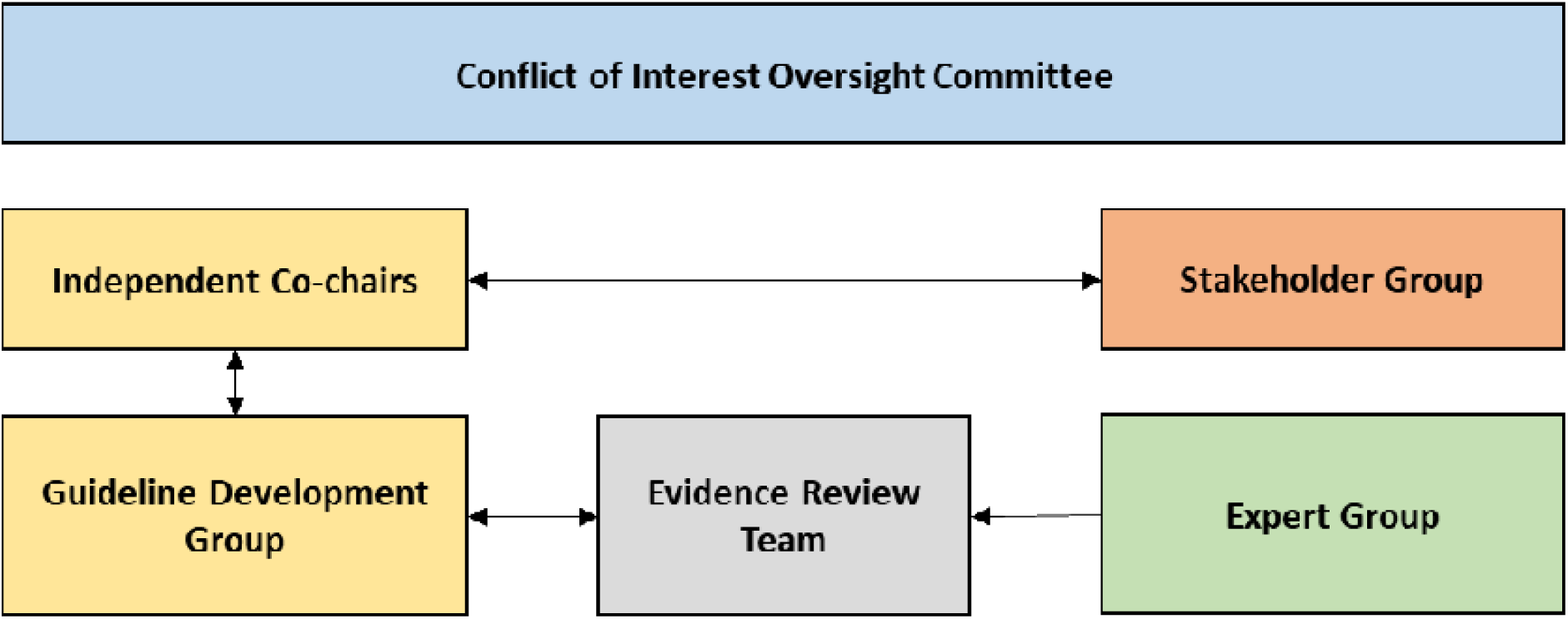
Guideline governance structure.

#### 2.1.1 Guideline Development Group

The GDG comprises members with clinical (e.g., general practice, nursing, pharmacy, psychiatry, psychology, psychotherapy), health economics, knowledge translation, mental health policy, methodological, neuroscience, pharmacology, and legal expertise, and lived experience. GDG members will be offered training in Aboriginal and Torres Strait Islander cultural awareness provided by the Australian Indigenous Doctors’ Association (Australian Indigenous Doctors’ Association) or Royal Australian College of General Practitioners (Royal Australian College of General Practitioners). GDG members will also be offered the opportunity to undertake We Al-li Trauma-informed Aboriginal and Torres Strait Islander Cultural Capability training (We Al-li). The GDG will formulate recommendations and good practice statements using the GRADE Evidence-to-Decision framework (refer to Methodology for Developing Recommendations section for details). Each GDG member will complete a declaration of interests form, which will be assessed by the COI Oversight Committee to ensure that members are not conflicted or that conflicts are managed according to a management plan in compliance with NHMRC requirements.

The GDG is co-chaired by a pharmacist and guideline methodologist with no direct or indirect financial interactions with any companies (or their foundation) involved in the development, production, or delivery of MDMA, another psychedelic, psychedelic-like drug, or related services. In line with NHMRC requirements, the co-chairs were appointed based on their independence and expertise in chairing and facilitating guideline meetings using GRADE methodology (National Health and Medical Research Council, 2018). Having two co-chairs will help maintain balance and independence. Both co-chairs have experience in leading or guiding the development of NHMRC-approved guidelines. The co-chairs will lead GDG and Stakeholder Group meetings, promote balanced participation of group members, and facilitate best practice application of the GRADE process. The guideline methodologist has a non-voting role, and no direct or indirect financial or non-financial interests, so will take primary responsibility for ensuring compliance with the conflict of interest policy.

#### 2.1.2 Stakeholder Group

The Stakeholder Group will comprise government agencies (e.g., Department of Veterans’ Affairs), professional organisations (e.g., Australian Psychological Society, Pharmaceutical Society of Australia, Royal Australian College of General Practitioners, Australian Indigenous Psychologists Association, Psychotherapy and Counselling Federation of Australia), peak bodies or research organisations (e.g., Australians for Mental Health Limited, Phoenix Australia, Black Dog Institute), and consumer representatives (e.g., Fearless, Arafmi, Tandem Carers, LGBTIQ+ Health Australia, Victorian Aboriginal Community Controlled Health Organisation, Heroic Hearts Project Australia, Indigenous Psychedelic-Assisted Therapies). The Stakeholder Group will provide strategic advice on the project, advise on barriers and risks, be invited to provide feedback during the public consultation process, and assist in disseminating the Guideline. Organisations represented on the Stakeholder Group may be invited to endorse the Guideline.

#### 2.1.3 Expert Group

The Expert Group will comprise authorised prescribers, consumers and carers, general practitioners, psychiatrists, psychologists, social workers, researchers, and others with relevant expertise. This includes but is not limited to, clinicians and consumers with direct experience providing and receiving MDMA-AP but who could not be part of the GDG due to involvement in industry-sponsored psychedelic clinical trials, private clinics, or other COIs. The Expert Group will not be a decision-making body but exists to provide input without compromising the actual or perceived integrity of the recommendation process. The Expert Group will not meet and discuss together as a group. The perspectives from the Expert Group will be solicited via one-on-one interviews (or, in the case of consumers, one-on-two interviews) facilitated by the Project Manager. The interviews will be audiotaped, transcribed *verbatim* and thematically analysed following the process outlined by Braun and Clarke (2021). Ethics approval has been obtained from the Monash University Human Research Ethics Committee for the in-depth interviews (Project ID: 44868). The interview findings will be presented to the GDG by a member of the ERT. A member of the COI Oversight Committee will review the material to be presented in advance of the meeting to ensure that the GDG is made aware of the COIs of the Expert Group. With the exception of the co-chairs and ERT, the GDG will not have direct contact in relation to the Guideline with the Expert Group members. Communication between the GDG and Expert Group relevant to the guidelines will be via the Project Manager.

#### 2.1.4 Conflict of Interest Oversight Committee

The COI Oversight Committee includes two guideline methodologists with no direct or indirect interests relevant to the Guideline topic. The COI Oversight Committee will advise on the COI policy and process, review the declarations of prospective GDG candidates, evaluate the interests of each GDG member, assess whether an interest reflects a conflict, and devise appropriate conflict management plans. A COI will be defined in accordance with the Australian Code for the Responsible Conduct of Research as a ‘situation where an independent observer might reasonably conclude that the professional actions of a person are or may be unduly influenced by other interests’ (Australia Universities, 2018). Potential COIs will include financial, organisational, or intellectual interests (refer to Supplementary material 1 for the COI Policy). All potential GDG members were asked to complete a Declaration of Interests Form prior to appointment. Disclosed interests will be updated and reviewed by the COI Oversight Committee prior to the start of each GDG meeting. Each disclosed interest will be individually assessed for its level of conflict risk based on the COI matrix adapted from the Clinical Guidelines Committee of the American College of Physicians (Qaseem et al., 2019) (Supplementary material 1). The COI Oversight Committee will notify the members of their management plans before the start of each meeting. Individuals with interests assessed as serious COIs will not be appointed to the GDG but may be invited to join the Expert Group. Members with moderate-risk COIs may contribute to discussions and be permitted to vote independently ahead of meetings. This vote will only be used to elicit the perspectives of members of the GDG and facilitate discussion. Members with a moderate-risk will not be permitted to vote in the meeting on either the judgements required in the GRADE Evidence-to-Decision framework or the recommendations. Members with low-risk COIs will be permitted to contribute to discussion and voting without restriction.

#### 2.1.5 Evidence Review Team

The ERT comprises the Project Manager and researchers with experience in evidence synthesis. The ERT will collate evidence on benefits and harms that the GDG will use to formulate recommendations and good practice statements. The ERT will provide evidence to support the GDG’s decision-making and may provide clarifying information during the GDG meetings, but will not contribute to the decision-making process itself. The ERT will conduct literature searches, formulate an overview of systematic reviews, and extract relevant data.

#### 2.1.6 Funder

The Guideline is funded by a philanthropic donation to the Faculty of Pharmacy and Pharmaceutical Sciences, Monash University. Funding was also received from Monash University, University of Melbourne, and The Florey Institute of Neuroscience and Mental Health. The philanthropic donor had no role in the design, data collection and analysis, decision to publish, or preparation of the Guideline, and they will not benefit materially from the outcomes of their funding. The philanthropic donor will be requested to sign a declaration stating that they have no potential COIs including financial, organisational, or intellectual interests, and that they will not have influence over the Guideline’s processes and outputs.

### 2.2 Methodology for Evidence Synthesis

The GRADE Evidence-to-Decision framework will be used by the GDG to make recommendations from the evidence and other information (Alonso-Coello et al., 2017; Alonso-Coello et al., 2016). The criteria considered in the GRADE Evidence-to-Decision framework are (1) benefits and harms, (2) certainty of the evidence, (3) values and preferences, (4) resources, (5) equity, (6) acceptability, and (7) feasibility (Alonso-Coello et al., 2017; Alonso-Coello et al., 2016).

#### 2.2.1 Question to be Addressed in the Guideline

The Guideline will address a single clinical question “In people living with PTSD, what are the benefits and harms of MDMA-AP compared to other or no treatment?” This Guideline will focus on MDMA because MDMA is at a more advanced stage of clinical development than psilocybin. MDMA has been the subject of two Phase 3 clinical trials (Mitchell et al., 2021; Mitchell et al., 2023).

#### 2.2.2 Criteria for Considering Studies for the Evidence Review

A hierarchical and staged approach will be used to identify and synthesise evidence about the effects of MDMA-AP on prioritised outcomes (benefits and harms) (Figure 2). The Participants, Interventions, Comparators, and Outcomes (PICO) eligible for inclusion in the evidence review are as follows.

1. Population: people living with PTSD. Specific populations with possible different treatment needs and outcomes will be considered while making recommendations, such as Aboriginal and Torres Strait Islander peoples, military and ex-military personnel, emergency services personnel, adolescents, older people, culturally and linguistically diverse peoples, neurodivergent individuals such as people with Autism Spectrum Disorder (ASD) or Attention Deficit Hyperactivity Disorder (ADHD), people with mental health co-morbidities, and people with different trauma types.
2. Intervention: MDMA-AP. The intervention was defined as MDMA-AP because the TGA approval does not include the use of MDMA as a standalone therapy and MDMA is typically combined with psychotherapy components. MDMA-AP reported in Phase 3 clinical trials has comprised three sessions of preparation, three sessions of experimental or dosing, and three sessions of integration following each experimental session. The psychotherapy component in the clinical trials – the “inner-directed” method has been described as a “non-directive approach that pertains to inviting inquiry and providing suggestion rather than directing the participant in the therapeutic approach”. The psychotherapy component itself is not a treatment option recommended in current PTSD treatment guidelines, such as the Australian Guidelines for the Treatment of Acute Stress Disorder, Posttraumatic Stress Disorder, and Complex PTSD (Phoenix Australia, 2020) and Effective Treatments for PTSD: Practice Guidelines from the International Society for Traumatic Stress Studies (Bisson et al., 2019; International Society for Traumatic Stress Studies, 2018).
3. Comparators: Other or no treatment. For the purpose of the evidence review, the comparator was deliberately defined broadly to include inactive comparators (e.g. placebo) and all types of treatment, such as psychological interventions, pharmacological interventions, watchful waiting, or no treatment. This broad definition was deemed necessary due to several challenges: the lack of evidence and treatment protocol for emerging treatments, inconsistent adoption of current guidelines by practitioners, limited accessibility of recommended treatments for certain populations (e.g., those in regional or remote areas), and issues of non-tolerance to exposure-based treatments. The broad definition of comparator accounted for clinical trials that have used active or inactive placebos together with psychotherapy. Active placebos have included low-dose MDMA in Phase 2 MDMA trials to mimic some of the physiological or psychological effects of MDMA without providing its full therapeutic benefits. Evidence from trials using an active placebo will be considered because active placebos may improve blinding and inclusion of these trials will maximise data availability. For safety outcomes, data from comparisons with an inactive placebo will be prioritised to avoid underestimation of potential MDMA-related adverse events.
4. Outcomes: Outcomes were determined through an outcome prioritisation process involving the GDG (see next) and included PTSD symptoms/diagnosis (e.g., symptom severity, remission, loss of diagnosis), co-occurring mental health symptoms, other patient-reported outcome measures (e.g., daily functioning, health-related quality of life), and adverse events (e.g., adverse drug events, suicide risk, MDMA diversion or misuse, treatment related discontinuity/ withdrawal, treatment burden).
5. Types of studies: systematic reviews of randomised trials (for effectiveness outcomes) and non-randomised studies (for safety outcomes only). Supplemental primary studies will be considered if meta-analyses for prioritised outcomes require updating (i.e. to include new or missing studies).

**Figure 2.**
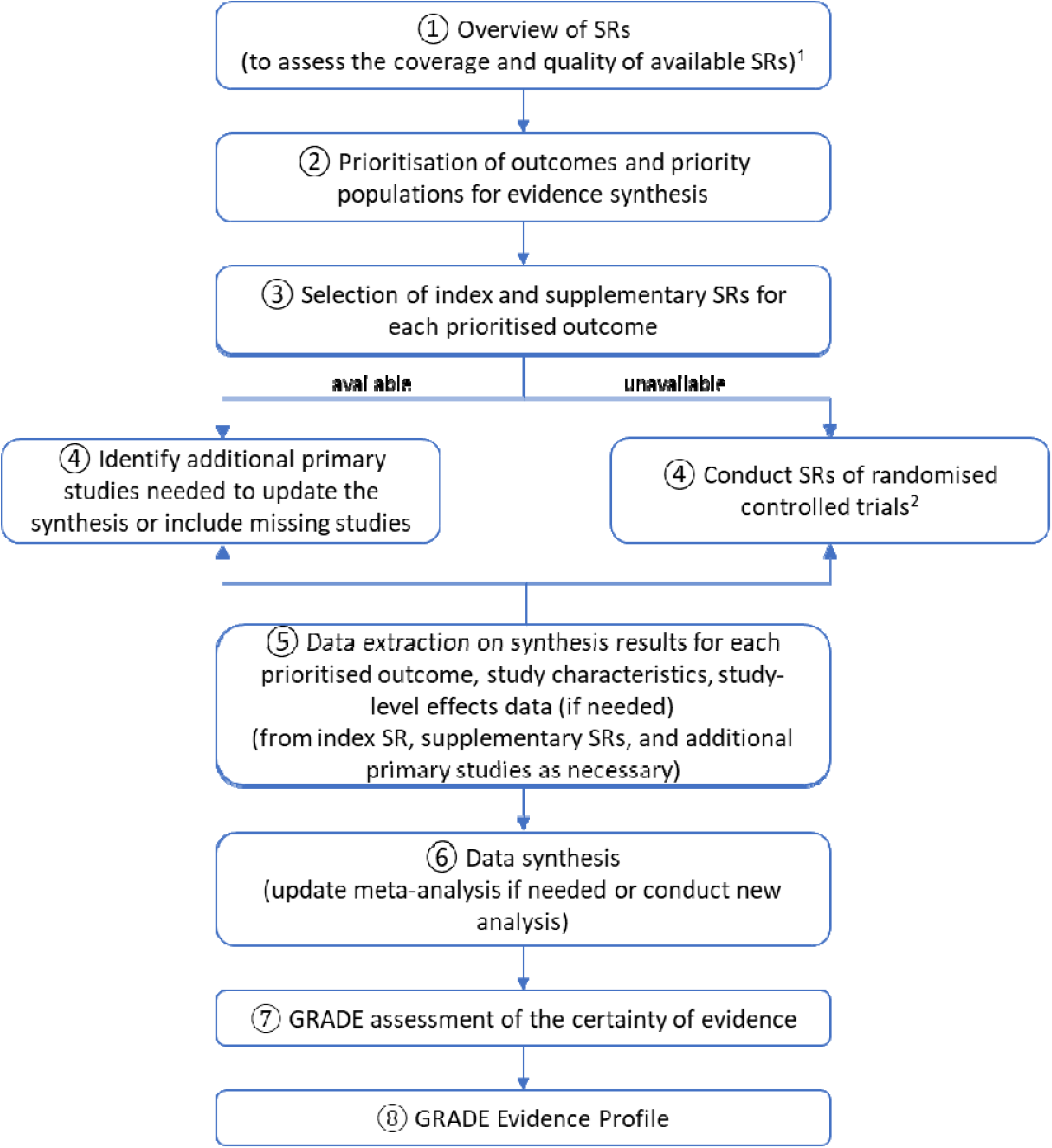
The process of synthesising evidence for benefits and harms. SR, systematic review ^1^Manuscipt has been published in a peer-reviewed journal (Yong et al., 2025). ^2^This was planned but the step was not needed because high-quality systematic reviews that met the PICO criteria were identified.

#### 2.2.3 Overview of Systematic Reviews

An overview of systematic reviews or meta-analyses was conducted to critically evaluate published and unpublished systematic reviews and meta-analyses relevant to the clinical question (Yong et al., 2025) (Figure 2, Step 1). The overview included a systematic search for reviews using keywords and Medical Subject Headings related to “MDMA”, “PTSD”, and “systematic review or meta-analysis” without any date or language restrictions. A grey literature search was conducted using the exact keywords to identify government reports, conference proceedings, or unpublished reports. The literature search and abstract screening were conducted independently by two reviewers. Any disagreements were resolved by discussion or involvement of a third reviewer when necessary. The lists of primary studies included in each review were compared and the degree of overlap was estimated using the corrected covered area (CCA) method proposed by Pieper et al. (2014). The methodological quality of the included reviews was assessed independently by two reviewers using the A MeaSurement Tool to Assess Systematic Reviews 2 (AMSTAR-2) tool (Shea et al., 2017).

#### 2.2.4 Rating Outcome Importance

GDG members rated the importance of specific outcomes for decision-making (Figure 2, Step 2). Potentially relevant outcomes were identified from the outcomes prioritised in existing PTSD guidelines (Bisson et al., 2019; International Society for Traumatic Stress Studies, 2018; Phoenix Australia, 2020), outcomes measured in systematic reviews or meta-analyses of psychological and pharmacological interventions for PTSD (Coventry et al., 2020; Karatzias et al., 2019; Mavranezouli et al., 2020), and safety outcomes raised in the US FDA’s Psychopharmacologic Drugs Advisory Committee Meeting with Lykos Therapeutics (Food and Drug Administration, 2024). Of the 13 prioritised outcomes, eight were rated as “critical” (mean score of 7 to 9), including suicide risk, change in PTSD symptoms, daily functioning, health-related quality of life, change in self-organisation/emotional regulation, change in depressive symptoms, cardiac-related adverse events, impact on productivity, and change in anxiety symptoms (Supplementary material 2). Five outcomes were rated as “important” (mean score of 4 to 6), including change in anxiety symptoms, treatment discontinuation/withdrawal, adverse events, sleep quality, and diversion or misuse of MDMA. It was noted that GDG members showed diverse opinions regarding the importance of the proposed outcomes. Of the 13 outcomes presented, 11 were selected by at least one GDG member as being among the top three most important outcomes for the Guideline, except change in anxiety symptoms and treatment discontinuation. Since all 13 proposed outcomes were rated as either critical or important for decision-making, they will all be included in the Summary of Findings.

GDG members also identified several critical outcomes not covered in the list (Supplementary material 2). These outcomes will be considered in the Evidence-to-Decision framework. For each prioritised outcome, the GDG discussed the minimal important difference for each outcome (Guyatt et al., 2011; Neumann et al., 2022; Zeng et al., 2021).

#### 2.2.5 Selection of Systematic Reviews and Additional Studies for Informing the Guideline

High-quality (AMSTAR-2 overall rating as high) systematic reviews identified in the overview of systematic reviews may be used as index systematic reviews (for data extraction into the evidence profile) or as supplementary reviews (for additional data needed to address missing studies or apply the GRADE Evidence-to-Decision framework) (Figure 2, Step 3). Four systematic reviews were identified in the overview of systematic reviews: Colcott et al. (2024); Heath et al. (2022); Kisely et al. (2023); Mustafa et al. (2024).

If the literature search in an index systematic review used to generate the evidence profile is more than 12 months old when the draft Guideline is released for public consultation, the literature search will be updated to identify any new studies (Figure 2, Step 4). It was not deemed necessary to conduct a new systematic review because high quality systematic reviews meeting the PICO criteria were identified.

#### 2.2.6 Data Extraction

For each prioritised outcome, data will be extracted for each primary study from the index systematic review, supplementary systematic reviews and primary studies (if needed) by one reviewer and checked by another (Figure 2, Step 5). From the index systematic reviews, the following information will be extracted: characteristics of each trial (trial PICO, participant characteristics, risk of bias, number of participants randomised), outcomes measured, measurement methods, measurement timepoints, results data (meta-analytic effect estimates for each prioritised outcome, number of studies and participants contributing to each analysis, outcome measures in mean difference, and details required for interpretation), and other data needed to assess certainty of evidence (such as assessment of bias due to missing results from the synthesis, sensitivity analyses, subgroup analyses, and data needed to assess inconsistency such as forest plots). If comparison across systematic reviews indicates that a study is missing from the meta-analyses in the index systematic review, primary study data will be extracted from a supplementary systematic review and the meta-analyses will be updated. If a prioritised outcome is not addressed in any of the systematic reviews, the data will be extracted from relevant primary studies and relevant trial registries and reports will be checked to identify if the outcome has been measured or reported. Primary studies will also be used to extract other missing information or where there are unexplained discrepancies between systematic reviews. The quality of additional primary studies identified from an updated search will be assessed using the Cochrane risk-of-bias tool for randomised trials (RoB 2) (Higgins et al., 2019).

#### 2.2.7 Data Synthesis

Meta-analyses identified from existing reviews will be updated if needed, such as to add or remove studies or change the grouping within the analyses (Figure 2, Step 6). When an update is needed, the summary statistics of each study will be extracted from the forest plots or tables of the index systematic review to be pooled with that from additional primary studies.

New meta-analyses will be conducted if none have been identified. Mean differences will be calculated if all included studies use the same scale, whereas standardised mean differences will be used for studies with different scales. Binary outcomes will be estimated using relative risks or hazard ratios. Meta-analyses of the subgroups will be conducted if there is sufficient homogenous population. The subgroups will be prioritised by the GDG ahead of synthesis. If necessary, sensitivity analyses will be undertaken to assess the impact of low-quality or withdrawn trials on the overall findings. If a meta-analysis is not possible, the results will be presented narratively.

#### 2.2.8 GRADE Assessment of the Certainty of Evidence

The certainty of evidence for all critical and important outcomes will be assessed using the GRADE approach based on five GRADE domains: risk of bias, imprecision, inconsistency, indirectness, and publication bias (Figure 2, Step 7) (Balshem et al., 2011).

#### 2.2.9 GRADE Evidence Profile

Evidence for benefits and harms will be presented in a GRADE Evidence Profile. This profile will include all outcomes considered to be critical and important for making a recommendation, results for each outcome (reported as the absolute and relative magnitude of effect), number of participants and studies, overall certainty of evidence for each outcome, and a plain language summary to convey the evidence findings based on GRADE informative statements (Figure 2, Step 8) (Santesso et al., 2020).

#### 2.2.10 Evidence-to-Decision Framework

Evidence for domains 1 and 2 (benefits and harms, certainty of the evidence) will be derived from the GRADE Evidence Profile. Narrative literature reviews will be conducted to identify evidence for domains 3 to 7 (values and preferences, resources, equity, acceptability, and feasibility). Summary of findings from the semi-structured interviews with each member of the Expert Group that are relevant to the framework will be presented to the GDG. Input derived from the discussions among the GDG members will also be captured.

### 2.3 Development of Recommendations

The Evidence Profile and findings from the narrative review and in-depth interviews will be presented to the GDG at a series of GDG meetings. Prior to group discussion, each GDG member will independently judge whether the evidence and other information for each domain in the Evidence-to-Decision framework support a recommendation for the intervention (MDMA-AP) or the alternative option. The summary judgements for each domain are shown in Table 1. Anonymous voting will be conducted using electronic polls. This will ensure that the views and perspectives of each member are captured without the influence of other members. The Methodology Chair will then facilitate a group discussion about each domain to explore possible differences in voting. The voting process will be repeated after the group discussion.

**Table 1.**
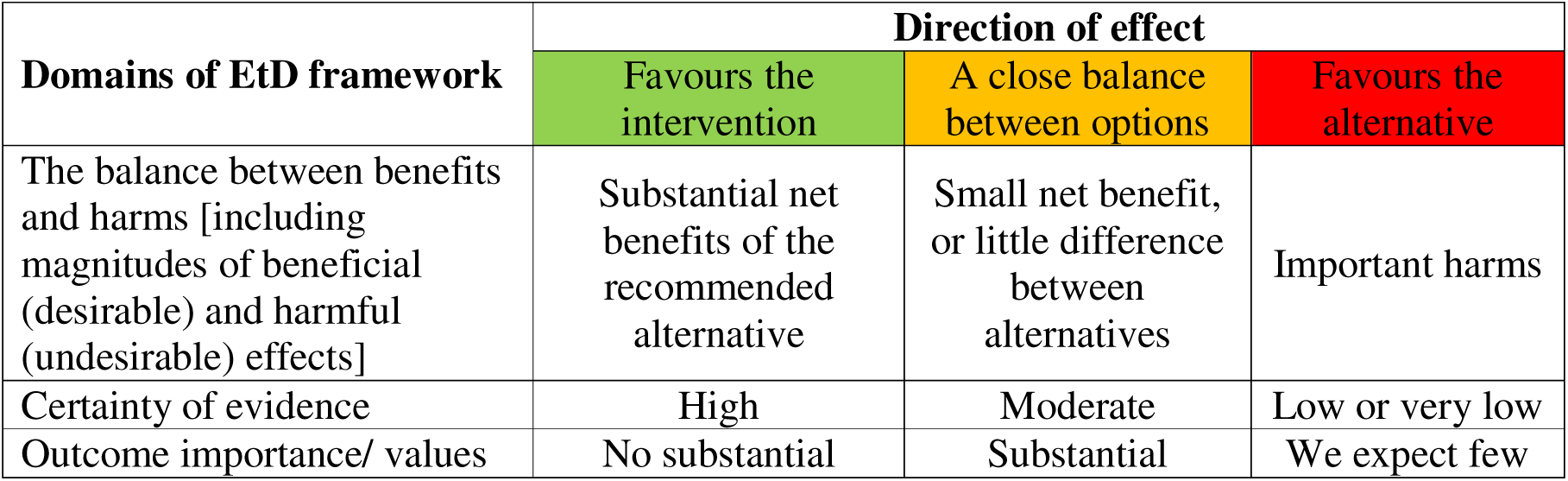
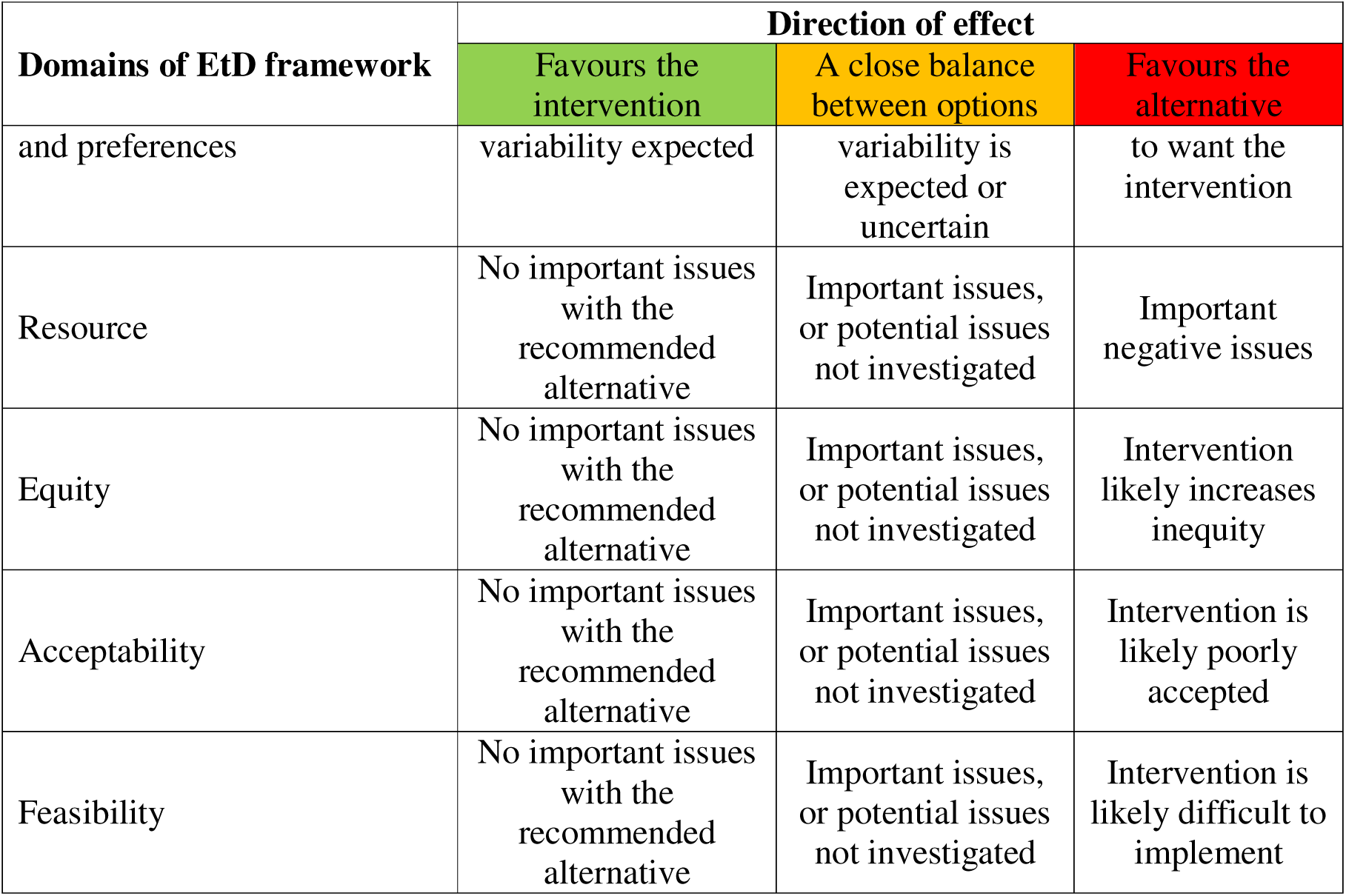
Phrasing implemented in MAGICapp to express the GDG judgement of whether the evidence and other information for each Evidence-to-Decision (EtD) factor supports a recommendation for the intervention or the alternative option.

| Domains of EtD framework | Direction of effect |  |  |
| --- | --- | --- | --- |
|  | Favours the intervention | A close balance between options | Favours the alternative |
| The balance between benefits and harms [including magnitudes of beneficial (desirable) and harmful (undesirable) effects] | Substantial net benefits of the recommended alternative | Small net benefit, or little difference between alternatives | Important harms |
| Certainty of evidence | High | Moderate | Low or very low |
| Outcome importance/ values | No substantial | Substantial | We expect few |
|  | Favours the intervention | A close balance between options | Favours the alternative |
| and preferences | variability expected | variability is expected or uncertain | to want the intervention |
| Resource | No important issues with the recommended alternative | Important issues, or potential issues not investigated | Important negative issues |
| Equity | No important issues with the recommended alternative | Important issues, or potential issues not investigated | Intervention likely increases inequity |
| Acceptability | No important issues with the recommended alternative | Important issues, or potential issues not investigated | Intervention is likely poorly accepted |
| Feasibility | No important issues with the recommended alternative | Important issues, or potential issues not investigated | Intervention is likely difficult to implement |

After voting on each domain within the Evidence-to-Decision framework has been completed, the GDG will vote on the direction (for or against MDMA-AP) and strength (strong or conditional) of the overall recommendation (Table 2) (Andrews et al., 2013). Where possible, voting on the overall recommendation will be by unanimous agreement or consent by all members. If unanimity cannot be achieved, the group will discuss the concerns and reservations, which may lead to modification or rewording of the recommendation. If unanimity is still not reached after discussion, a majority vote will be sought. The reasons for disagreement or objection of members will be documented in the Technical Report.

**Table 2.** Types of possible recommendations and their definitions based on GRADE Handbook.

| Recommendation | Definition |
| --- | --- |
| Strong Recommendation for | The GDG is confident that the benefits of an intervention outweigh its harms. The intervention will likely benefit most or all individuals. |
| Strong Recommendation against | The GDG is confident that the harms of an intervention outweigh its benefits. The intervention will likely not benefit most or all individuals. |

| **Recommendation** | **Definition** |
| --- | --- |
| Conditional Recommendation for | The benefits probably outweigh the harms, but appreciable uncertainty exists. The intervention may not be suitable for all individuals. There is a need to carefully consider the individual patient's circumstances, preferences, and values. |
| Conditional Recommendation against | The harms probably outweigh the benefits, but appreciable uncertainty exists. The intervention may not be suitable for all individuals. There is a need to carefully consider the individual patient's circumstances, preferences, and values. |
| Recommendations to use interventions only in research | There is to date insufficient evidence to support a decision for or against an intervention. Further research has large potential for reducing uncertainty about the effects of the intervention. Further research is considered to be of good value for the anticipated costs. |

### 2.4 Development of Good Practice Statements

A series of Good Practice Statements related to principles important to follow prior to, during, and after the initiation of MDMA-AP for PTSD will be formulated if there is clear indirect evidence that their implementation will provide unequivocal benefits. Good Practice Statements are actionable statements grounded in either: (1) ethical principles or human rights (e.g., the right to health and to make decisions about treatment with informed consent), (2) essential principles, practices, and protocols (e.g., to guide standards for infection control), or (3) established scientific evidence. The Evidence-to-Decision criteria will be considered for each Good Practice Statement to decide if there is high certainty that the desirable effects outweigh undesirable effects or that the opposite course of action would be clearly inappropriate (Dewidar et al., 2023). All recommendations and Good Practice Statements will be discussed, drafted, and approved by the GDG. These recommendations and Good Practice Statements will not require endorsement by Monash University, the funders, or any of the other GDG members organisations in order to be published.

### 2.5 Dissemination and Implementation

The Guideline will be published on the digital platform MAGICapp and in peer-reviewed publications. The findings will be disseminated to medical and allied health professionals via digital dissemination, professional networks, conferences, and member organisations of the Stakeholder Group. A Consumer Companion Guide will be developed and disseminated to people living with PTSD and their carers, family members, and other supports.

## 3. Discussion

This protocol outlines a governance structure designed to uphold transparency, independence, and objectivity in accordance with NHMRC standards. The recommendations in the Guideline will be formulated based on a systematic process that considers evidence on benefits and harms in the context of the local health system and clinical practice.

The Guideline will be developed by utilising an integrated knowledge translation approach, emphasising the co-production of knowledge through active participation and shared decision-making between researchers and end-users (Gagliardi et al., 2017; Graham & Tetroe, 2009; Jull et al., 2017). Early engagement and sustained collaboration have been shown to enhance the uptake and implementation of research recommendations significantly (Gagliardi et al., 2016; Kothari & Wathen, 2013). For the context of this Guideline, the integrated knowledge translation approach enables the researchers and guideline developers to better understand the practice environment, interpret results within their contextual framework, and enhance the real-world applicability of the Guideline. At the same time, end-users gain a clearer understanding and awareness of the research process and are given opportunities to highlight their clinical needs. The in-depth interviews with the Expert Group will be conducted as part of the integrated knowledge translation process to ensure that the perspectives of end-users are incorporated throughout the guideline development. The Stakeholder Group, comprising key government agencies, non-government organisations, and professional organisations, will be convened throughout the Guideline development to advise implementation barriers and sensitivities and identify implementation strategies. Perspectives of different populations will be considered, such as those in non-metropolitan settings (rural and remote), Aboriginal and Torres Strait Islander peoples, culturally and linguistically diverse peoples, neurodivergent individuals, and LGBTQIA+ communities.

Given the nascent stage of psychedelic research, it is anticipated that the GDG and Expert Group will include some members with financial, organisational or intellectual COIs. Therefore, measures have been taken to ensure decisions are independent and objective. These measures include (1) ensuring diversity of perspectives and interests among members, (2) requiring GDG and Expert Group members to declare relevant interests, (3) allowing GDG members to vote on the Evidence-to-Decision framework independently before group discussion, and (4) forming an Expert Group to provide input while remaining independent from the GDG. These measures ensure perceived COIs are identified and managed, thereby reducing potential concerns about objectivity and increasing public trust (National Health and Medical Research Council, 2019; Schünemann et al., 2015).

In conclusion, the development of an Australian clinical practice guideline for MDMA-AP in PTSD will provide clinicians with evidence-based recommendations to advance the field of psychedelic-assisted therapy. The Guideline will involve input from a broad clinical experts, researchers, and consumers to help ensure appropriate use of MDMA-AP.

## Data Availability

Details of the current protocol are included within the manuscript and supplementary material.

