## Supplementary Material for "Development of an Australian Clinical Practice Guideline on methylenedioxymethamphetamine (MDMA)-assisted Psychotherapy for Post-traumatic Stress Disorder"

### Supplementary Material 1. Conflict of Interest Policy

Conflict of Interest Policy:

Clinical Practice Guidelines on the Appropriate Use of Methylenedioxymethamphetamine (MDMA)- assisted Psychotherapy for Post-traumatic Stress Disorder

### **Purpose**

All individuals involved in the Guideline Development Group (GDG) are required to complete a Declaration of Interests Form. If a conflict of interest (COI) is identified, it will be managed in accordance with the National Health and Medical Research Council (NHMRC) policies and procedures. According to the NHMRC Disclosure of interests and management of conflict of interest guide and under the *Australian Code for the Responsible Conduct of Research*:

‘*COI exists in a situation where an independent observer might reasonably conclude that the professional actions of a person are or maybe unduly influenced by the other interests. The perception that a COI can raise concerns about the integrity of the individuals or the management practices of the institution, potentially undermining community trust in research.’*

### **Scope**

This COI policy applies to all individuals participating in the decision-making processes relating to the development of recommendations and good practice statements related to MDMA-assisted psychotherapy for PTSD. These recommendations and good practice statements will be incorporated in new Clinical Practice Guidelines on the Appropriate Use of Psychedelics in Mental Health Conditions. This COI policy is a living document and updates and edits may occur in real time.

### **Definition of Psychedelics**

Psychedelics (also known as hallucinogens) are a class of psychoactive substances that produce changes in perception, mood and cognitive processes. Examples of psychedelics include psilocybin, ayahuasca, LSD, DMT, NBOMes, and mescaline. Methylenedioxymethamphetamine (MDMA) is classified as an empathogen or entactogen, but for simplicity, it will be referred to as a psychedelic throughout this guideline. Cannabinoids (e.g., cannabis) and dissociatives (e.g., ketamine, nitrous oxide) are not classified as psychedelics within the scope of this guideline.

### **Types of Conflict of Interests**

According to the NHMRC’s Disclosure of interests and management of conflict of interest: A guide supporting the *Australian Code for the Responsible Conduct of Research*:

‘Financial interests are the foremost in the public mind, but other interests may also be relevant, including personal, familial, professional and organisational.’

Financial interests requiring disclosure include, but are not limited to:

- direct payment, such as salary or consultancy fees;
- indirect payments, for example funding of travel or accommodation
- payments to support research, such as funding from an industry or interest group;
- company shares or options;
- royalties;
- directorships;
- some scholarships; and
- operational or infrastructure support.

Disclosure may also be required when a financial interest of the kind listed above is held by an immediate family member. Financial interests also exist where there is a future expectation of a benefit, for example, proceeds from the sale of intellectual property arising from a project or the promise of shares in a spin off company.

Non-financial interests that require disclosure include, but are not limited to:

- represent or have roles/ affiliations with an organisation that could stand to benefit from or be affected by the guideline (e.g., advocacy group, industry, industry-linked foundation);
- personal or social relationships and current and past professional relationships, where relevant;
- recent employment with, or role in, organisations with financial links or affiliations with industry groups that could stand to benefit from or be affected by the guideline; and
- intellectual interests related to psychedelics
- specific questions adapted from the US Preventive Services Task Force Disclosure Form will also be included to understand the member’s stance or views related to psychedelics (see table below)

| *To the best of your ability, please respond ‘yes’ or ‘no’ to each of the questions below. If the answer is ‘yes’ for any question, please include details or references that may be helpful in evaluation the potential influence of each relationship or personal belief. A ‘yes’ answer will not necessarily disqualify you from participating in the Guideline.* | | |
| --- | --- | --- |
|  | **Yes** | **No** |
| 1. Do you have strongly held beliefs/ perspectives related to psychedelic-assisted therapy? |  |  |
| *If ‘yes’, explain:* | | |
| 2. Would your beliefs/ perspectives (in question above) make it difficult for you to work in an unbiased manner on the Guideline? |  |  |
| *If ‘yes’, explain:* | | |
| 3. Have you ever authored, co-authored, or publicly provided an opinion related to psychedelic-assisted therapy? |  |  |
| *If ‘yes’, explain:* | | |
| 4. To the best of your knowledge, do you work for, or are you a member of, an organisation with a stated position (e.g., position statement, blog, editorial, legislature or legal testimony, or related document) related to this Guideline? |  |  |
| *If ‘yes’, explain:* | | |
| 5. Are you involved in formulating/ voting for positions in any organisation with a stated position related to the Guideline? |  |  |
| *If ‘yes’, explain:* | | |
| 6. Could this recommendation statement conflict with policies you have promoted or are obliged to follow? |  |  |
| *If ‘yes’, explain:* | | |

When disclosing interest, the COI Oversight Committee will consider the significance of the financial interest, including:

- the monetary value of the payment, gift or interest;
- whether payment was made to the individual directly, to their employer or to their immediate family;
- significance that a reasonable, independent observer would attach to the payment gift or interest;
- circumstances under which a gift or payment is made, for example is the gift or payment is regular payment or single instance; and
- potential influence or bias an affiliation may have on guideline development.

### **Process of Disclosure**

All contributors to decisions relating to guideline development are required to complete a Declaration of Interests Form that is to be returned to the Project Manager/Secretariat prior to attending their first meeting in which the development of recommendations is discussed.

- All actual, potential or perceived interests should be disclosed. Disclosure is required in relation to activities two years preceding and anticipated disbursements in the 12 months following appointment to GDG.
- It is the responsibility of individuals to continuously disclose any interest that is potentially a financial or non-financial COI. Members are expected to promptly inform the project manager/secretariat if there are any changes prior to or during the course of the guideline development.
- Members will be reminded of the need to disclose and update their declaration of interest at the start of each guideline development meeting.
- In line with the NHMRC requirements, failure to disclose a relevant interest, or knowingly misleading the group regarding potential conflicts of interest, could lead to sanctions such as removal from the GDG.

Completed Disclosure of Interest Forms and management plans will be kept electronically by the Project Manager/Secretariat. A register of disclosed interests will be recorded by the Project Manager/Secretariat throughout the duration of the project. The register is to be reviewed and endorsed by the COI oversight committee prior to the start of each GDG meeting. Members who are judged to have conflicts will be notified of the management plans (e.g., recuse from meeting, abstain from voting).

Conflicts of interest and corresponding management plans will be published for all members with voting power on recommendations.

### **Non-appointable Members**

Individuals who declare significant personal financial interests in one or more companies with a commercial interest in the outcomes of the guideline, and those with intellectual COIs that cannot be adequately managed will be excluded from the GDG. COIs that clearly preclude participation of an individual in a GDG include:

- Any involvement in industry-funded clinical trial(s) investigating the specific psychedelic under consideration in the guideline
- Any involvement in a broad portfolio of research funded primarily by the psychedelic industry
- Any involvement in private practice with potential income from the prescription or delivery of care related to psychedelics
- Any ownership of more than AUD$ 5000 worth of shares in a company that conducts research, manufactures or sells the psychedelic under consideration in the guideline
- Holding a patent on a product or technology that may be recommended in the guideline

Instead of being part of the GDG, these individuals will be invited to be part of the Expert Group if their input and expertise are deemed essential (refer Appendix A for the Terms of Reference for the Expert Group).

### **Conflict of Interest Identification**

Completed Declaration of Interest Forms will be reviewed by the Project Manager/Secretariat and a Conflict of Interest Oversight Committee comprising several guideline development team members to determine if any declared entries constitute a COI. If at any point in time a person is found to have purposely withheld or not disclosed information, that person may be removed from the GDG.

### **Conflict of Interest Management**

It is anticipated that the GDG will include some members with financial and/or intellectual conflicts of interest due to psychedelics being a new and niche research area. For this reason, efforts will be made to balance the perspectives of these individuals in the group. This includes:

- Ensuring diversity of GDG membership by selecting members with diverse perspectives, training, experiences, and interest towards psychedelics.
- Giving GDG members the opportunities to independently vote/ indicate their opinions on specific domains within the GRADE Evidence to Decision framework both prior to and during the meeting.

Each disclosed interest will be individually assessed for its level of conflict risk based on the matrix below. The matrix is adapted from the Clinical Guidelines Committee of the American College of Physicians (Qaseem et al., 2019) and ANZMUSC Guidelines Conflict of Interest Risk Assessment Method (Glennon, 2023).

|  |  | Interaction | | |
| --- | --- | --- | --- | --- |
|  |  | High risk^a^ | Moderate risk^b^ | Low risk^c^ |
| Entity | High Risk  (Pharma or other biomedical industry, insurers) |  |  |  |
|  | Moderate Risk (professional, consumer or advocacy bodies, foundations, non-industry-funders related to psychedelics) |  |  |  |
|  | Low Risk (Academic or educational institutions, publishers, government funders with no role in the research (e.g., MRFF, NHMRC) |  |  |  |

^a^High:

Active financial relationship, including direct payment to individual or family member (e.g. consulting, advisory boards, paid speaker, investments or equity), payment in kind (e.g. air travel for conference other than as invited speaker), non-academic publication (e.g. book deal)

Direct^[1]^ intellectual interest (i.e. research, academic activity, publication (e.g., journal article, textbook chapter), or advocacy directly relevant to recommendation topic)

Critical role in research directly or indirectly relevant to the recommendation topic (e.g. chief investigator, project initiator).

^b^Moderate:

Indirect^[2]^ intellectual interest (i.e. research, academic activity, publication (e.g., journal article, textbook chapter), or advocacy unrelated or indirectly relevant to recommendation topic)

Indirect financial relationship (e.g. work resulting in payment to hospital or university department, including investigator on industry-sponsored trial)

^c^Low:

Dissolved or historical financial relationship; minor financial relationship (e.g. attendance at educational dinner meeting); remote financial relationship (e.g. unrelated payment or contract with hospital or university department); other minor interaction (e.g. visits from pharmaceutical representative)

Personal experience

A management plan will be developed by the COI oversight committee to determine the degree to which a member may participate in the guideline development, based on the following principles:

- Members with high risk conflicts (‘red’) will be invited to join the Expert Group instead.
- Members with moderate risk conflicts (‘amber’) may contribute to the panel discussion, are permitted to vote independently using the GRADE EtD framework ahead of the meeting, but are not permitted to vote for the final recommendations.
- Members with low risk conflicts (‘green’) are permitted to contribute to discussion and voting without restriction.

The chair should have no financial interactions with any companies (or their foundation) that develops or produces MDMA, another psychedelic or psychedelic-like drug, for any indications. In the case of any disagreement about management plans, the plans will be reviewed by an independent Monash University staff member. An individual may elect to convert a high-risk conflict to a low risk conflict by immediately severing all financial ties. This action will be recorded by the secretariat.

**Appendix A**

### **Terms of Reference**

### *Expert Group*

1. Purpose:

The Expert Group (EG) is established to provide essential expertise and insights for the development of clinical practice guidelines. The primary aim of the EG is to ensure the practicality, relevance, and applicability of the guidelines to real-world clinical settings. The EG will have no role in making decisions or drafting recommendations relevant to the guidelines.

2. Composition:

The EG will consist of authorised prescribers, practitioners, researchers, advocacy groups, and consumers with clinical backgrounds and expertise relevant to the scope of the clinical practice guidelines under development.

3. Roles and Responsibilities:
a) Providing expert input: The EG may be invited to comment on information presented in the GRADE Evidence to Decision framework according to their expertise or the perspectives of the group they are representing. This may include but is not be limited to:
   - importance of outcomes (the value consumers place on main outcomes of the intervention)
   - resource requirements
   - impact on health equity
   - acceptability and feasibility of interventions (stakeholder perspectives)
   - contextualisation of guidelines within clinical pathways
b) Participating in qualitative interview: EG members will be interviewed individually to explore their perspectives on the management of MDMA in PTSD. The findings of the interview will be presented to GDG to be considered during decision-making. The findings might also be presented in conference or published in journals.
c) Ensuring practicality and relevance: EG members will ensure that the guidelines are practical and relevant to real-world clinical scenarios. Since the size of a GDG might limit the range of perspectives and experiences that can be accommodated on the GDG, having an EG helps address this through broader representation of individuals or groups with relevant experience or expertise.
4. Conflict of Interest Management:
a) The EG will not be involved in interpreting or grading the certainty of evidence, making judgements in relation to components of the Evidence to Decision framework, or making recommendations. Their role is strictly advisory, focusing on providing expert knowledge on the aforementioned components of the Evidence to Decision framework as relevant to their individual expertise.
b) That requests for input are framed as very specific questions, and advice is sought based on the role or area of expertise for which individuals are appointed to the EG.
c) The GDG will be advised to avoid compromising themselves through contact with EG members who could be perceived as influencing decisions. Any communications between the GDG and EG relevant to the guidelines should be diverted to the Secretariat/ project manager.
5. Amendments:

These Terms of Reference may be amended or updated as necessary to reflect changes in circumstances or to enhance the effectiveness of the EAG's operations. Any proposed amendments will be subject to review and approval by the conflict of interest oversight committee.

### Supplementary Material 2. Outcomes Importance as Rated by GDG Members

| **Outcomes identified from existing guidelines and systematic reviews** | **Mean** | **Min** | **Max** | **SD** |
| --- | --- | --- | --- | --- |
| ***Critical (mean score of 7-9)*** | | | | |
| Suicide risk (suicide, suicide attempts, self-harm associated with suicidal ideation, suicide ideation) | 8.15 | 7.00 | 9.00 | 0.77 |
| Change in PTSD symptoms | 8.00 | 7.00 | 9.00 | 0.88 |
| Daily functioning | 7.85 | 4.00 | 9.00 | 1.51 |
| Health-related quality of life | 7.38 | 6.00 | 9.00 | 1.00 |
| Change in self-organisation/ emotional regulation (eg, interpersonal problems, negative self-concept) | 7.23 | 6.00 | 9.00 | 0.97 |
| Change in depressive symptoms | 6.85 | 6.00 | 8.00 | 0.77 |
| Heart-related adverse events | 6.69 | 4.00 | 9.00 | 1.49 |
| Impact on productivity (e.g., employment and education) | 6.62 | 4.00 | 9.00 | 1.15 |
| ***Important (mean score of 4-6)*** | | | | |
| Change in anxiety symptoms | 6.46 | 5.00 | 8.00 | 0.93 |
| Treatment discontinuation/ withdrawal | 6.31 | 5.00 | 8.00 | 0.99 |
| Adverse events (e.g., Muscle tightness, decreased appetite, jaw clenching, excessive sweating, fatigue, restlessness, insomnia, nausea, blurred vision, chills) | 6.00 | 2.00 | 9.00 | 1.66 |
| Sleep quality | 6.00 | 2.00 | 8.00 | 1.62 |
| Diversion or misuse of MDMA | 5.38 | 1.00 | 9.00 | 2.43 |
| **Additional outcomes that were considered important by GDG members** | | | | |
| long-term outcomes (e.g., exacerbation of symptoms, sustained change in symptom severity) | | | | |
| comorbid symptoms (e.g., symptoms of numbing, problems with concentration, irritability) | | | | |
| change in quality of interpersonal relationships | | | | |
| alcohol and other substance misuse | | | | |

1. Same psychedelic and same indication (i.e. MDMA for PTSD), same psychedelic but different indication (e.g. MDMA for anxiety), different psychedelic but same indication (e.g. ayahuasca for PTSD).
2. Different psychedelic for different indication (e.g. psilocybin for depression, LSD for ADHD)
